# Prevalence and predictors of tuberculosis infection among people living with HIV in a high tuberculosis burden context

**DOI:** 10.1101/2022.12.04.22283086

**Authors:** Lilian N. Njagi, Videlis Nduba, Marianne Mureithi, Jared O. Mecha

## Abstract

**Background:** Tuberculosis (TB) disease is the leading cause of mortality among people living with the Human immunodeficiency virus (PLHIV). Interferon-gamma release assays (IGRAs) are approved for TB infection ascertainment. However, current IGRA data on the prevalence of TB infection in the context of near-universal access to antiretroviral therapy (ART) and widespread TB preventive therapy (TPT) implementation are lacking. We estimated the prevalence and determinants of TB infection among PLHIV within a high TB and HIV burden context.

**Methods:** This cross-sectional study included data from adult PLHIV age ≥ 18 years in whom QuantiFERON-TB Gold Plus (QFT-Plus) assay, an IGRA, was performed. TB infection was defined as a positive or indeterminate QFT-Plus test. Participants with TB and those who had previously used TPT were excluded. Regression analysis was performed to identify independent predictors of TB infection.

**Results:** Of 121 PLHIV with QFT-Plus test results, females were 74.4% (90/121), and the mean age was 38.4 (standard deviation [SD] 10.8) years. Overall, 47.9% (58/121) were classified as TB infection (QFT-Plus test positive and indeterminate results were 39.7% [48/121] and 8.3% [10/121], respectively), with mean ages of 38.7 (SD 10.30) vs 38.2 (SD 11.3) years, respectively (p=0.602). Being obese/overweight (body mass index ≥25; p=0.013, adjusted odds ratio (aOR) 2.90, 95% confidence interval [CI] 1.25–6.74) and ART usage for >3 years (p=0.013, aOR 3.99, 95% CI 1.55– 10.28) were independently associated with TB infection.

**Conclusion:** There was a high TB infection prevalence among PLHIV. A longer period of ART and obesity were independently associated with TB infection. The relationship between obesity/overweight and TB infection may be related to ART and immune reconstitution and requires further investigation. Given the known benefit of test-directed TPT among PLHIV never exposed to TPT, its clinical and cost implications for low and middle-income countries should be explored further.

**KEY MESSAGES:** *What is already known on this topic:* Among people living with HIV (PLHIV), the risk of progression to tuberculosis (TB) disease is higher with confirmed and untreated TB infection. Data on the prevalence of TB infection in the context of near-universal access to antiretroviral therapy (ART) and widespread TB preventive therapy (TPT) implementation are lacking in Africa.

*What this study adds:* This study provides evidence that the prevalence of TB infection remains high even in the context of near-universal ART and widespread TPT. ART use and obesity/overweight may be associated with TB infection.

*How this study might affect research, practice or policy:* This study should prompt larger studies to explore predictors of TB infection. TPT should remain as part of care for PLHIV on ART. A broader understanding of the clinical and cost implications of test-directed TPT for PLHIV in low and middle-income countries may better inform policy towards its recommendation.

## INTRODUCTION

Tuberculosis (TB) continues to be a global health concern. [1-3] Despite near-universal access to antiretroviral therapy (ART) and TB preventive therapy (TPT), TB remains the leading cause of disease and mortality among people living with HIV (PLHIV). [3-5] In 2021, approximately 10.6 million people developed TB worldwide, 6.7% (703 000) of whom were PLHIV. [3, 6] In the same period, 187,000 PLHIV died from TB, accounting for one-third of all AIDS-related deaths globally. [3, 5] The world health organisation (WHO) retained Kenya in the list of thirty high-burden countries for TB and HIV-associated TB for 2021–2025. [3] In 2021, the country had 133 000 incident TB cases, and 24% (32 000) were HIV/TB co-infected, of whom 34% (11 000) died. [6] TB infection, the precursor of TB disease, is a crucial focus for TB prevention strategies. [3, 7-9] The global burden of TB disease is estimated at a quarter of the population, [10] with wide regional variation. [11, 12]

Current information on the burden of TB infection in Africa and among PLHIV is sparse. [11, 12] There are also limited data on how the prevalence of TB infection has changed with near-universal access to ART and the widespread implementation of TPT. Other factors associated with TB infection among PLHIV are underexplored. [13-15] Establishing the burden of TB infection and the associated risk factors in PLHIV is important for various reasons. First, the risk of progression from TB infection to TB disease is higher among PLHIV with a positive test for TB infection than among those with a negative or unknown test result. [16] Second, TB risk is markedly reduced by TPT, with a 64% risk reduction among PLHIV with a positive test, compared to 14% among those with a negative or unknown test. [17] This has potential implications for practice given the country’s recommendation, supported by the WHO, to treat all PLHIV for TB infection without needing a confirmatory test. [9, 18-20]

The use of gold standard tests to establish disease burden is optimum when feasible. [21, 22] No gold standard exists for TB infection diagnosis. However, interferon-gamma release assays (IGRA) and tuberculin skin tests (TST) are screening tests approved by the WHO. [9] IGRAs are advantageous since they are not affected by prior BCG vaccination. [23] HIV impacts the QuantiFERON-TB Gold In-Tube test (QFT-GIT) less than it does the enzyme-linked immunospot assay (T SPOT-TB). [24, 25] The QuantiFERON-TB Gold Plus (QFT-Plus) and its forerunner, the QFT-GIT, have concordant performance. [26, 27] This paper presents the results of a study which sought to estimate the prevalence of TB infection among PLHIV using the QFT-Plus assay in a high TB burden context. Further, the determinants of TB infection in this population were explored.

## METHODS

### Study setting and design

This cross-sectional study used data collected at enrolment into a prospective observational study of adults aged ≥ 18 years from three HIV care and prevention centres in Nairobi, Kenya. The centres comprised Kenyatta National Hospital (KNH), Kenya’s largest national teaching and referral hospital; Pumwani maternity hospital, Kenya’s largest referral maternity hospital; and the Kenya Medical Research Institute, Centre for Respiratory disease Research (KEMRI CRDR), a centre of the regional leader in human health research. The clientele attending all three facilities is cosmopolitan, ranging from urban, peri-urban and rural settings.

### Study period and population

Participants seeking HIV care and prevention services from the three facilities between December 2019 and December 2020 were eligible to participate in the primary study. Verbal and written informed consent was obtained. All study participants underwent the recommended four TB symptom screening questions, including cough of any duration, fever, noticeable weight loss and night sweats. [28] Any participants with an affirmative response to any of the four questions underwent further evaluation, including a full physical examination, sputum examination for GeneXpert and chest x-ray to rule out TB disease. Participants suspected of having TB disease and those who had received isoniazid as a treatment for TB infection or disease were excluded from the study to reduce the chance of outcome misclassification (Figure 1).

**Figure 1.**
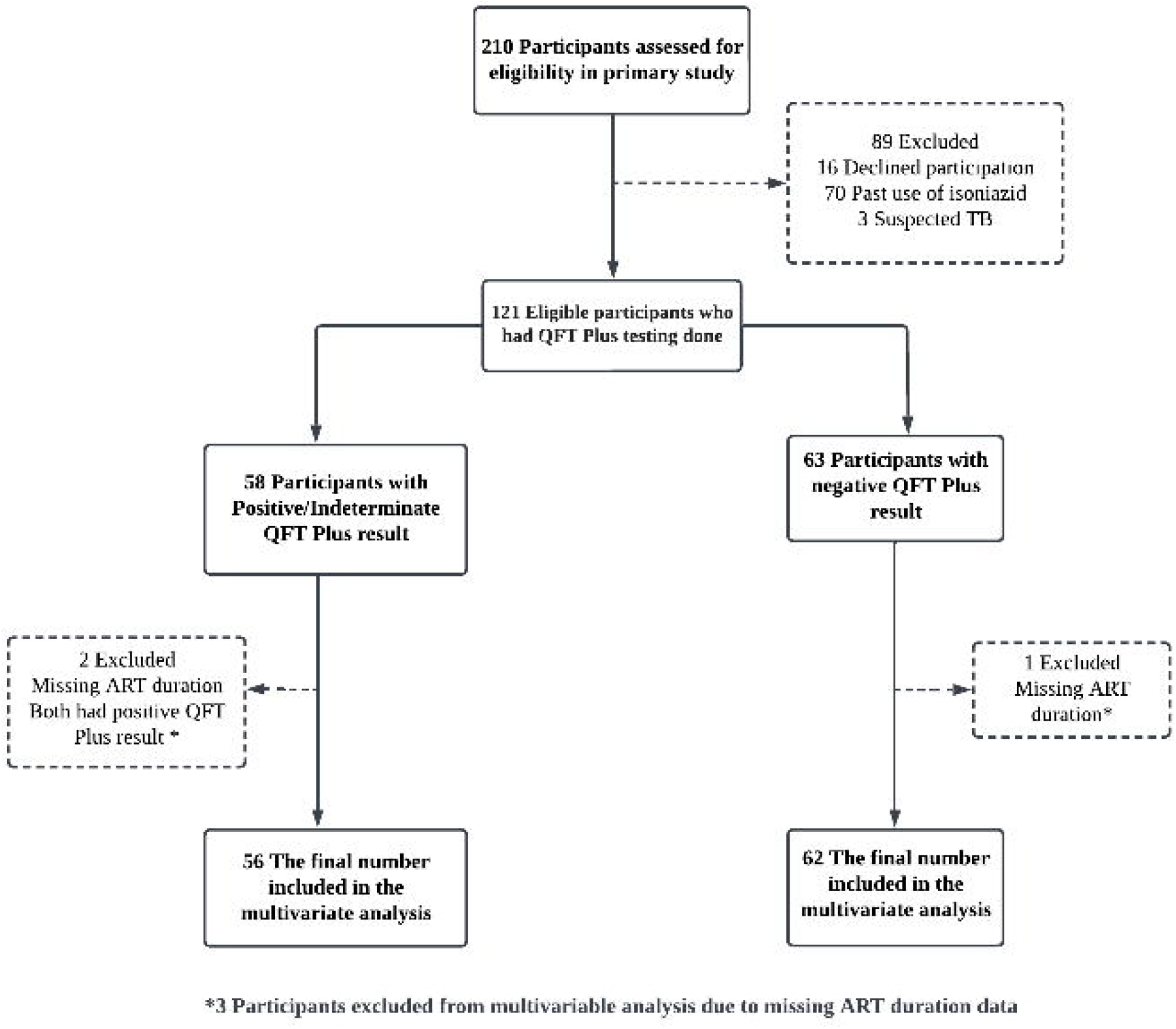
Enrolement and analysis flow diagram

### Variables and definitions

The outcome variable of interest was the TB infection status. TB infection was defined as a positive or indeterminate QFT-Plus result. We included indeterminate results in the definition of TB infection for several reasons. Foremost, we were bound by the existing WHO and country recommendations to treat all PLHIV for TB infection without requiring testing. [9, 18, 20, 28] Further, guidance on handling indeterminate QFT-Plus tests among PLHIV in the context of TPT is lacking. The WHO recommends a “case-by-case assessment for the potential benefit and harms of TPT” in regard to general testing for TB infection. [9] Second, the manufacturer suggests repeat testing for indeterminate results when related to technical factors where instructions are not followed. [29] In our case, instructions were followed per protocol. Finally, as this was a cross-sectional study and resources were limited, prospective repeat testing was impossible, and clinical care took precedence [30, 31]. All participants with negative results were considered not to have TB infection. The explanatory variables of interest were age, sex, body mass index (BMI) calculated from weight and height, cigarette smoking habit, alcohol use, household crowding, history of contact with a known TB case, diabetes status, duration of ART use as ≤3 versus >3 years, and viral load level as ≤ 40 versus > 40 HIV copies per millilitre of blood. To measure smoking and alcohol use habits, participants were asked if they had ever smoked and if they considered their alcohol use more than social. Similarly, participants were asked whether their household was crowded, whether they had been in contact with a person diagnosed with TB in the preceding two years, and if they had ever been diagnosed with diabetes.

### Clinical and laboratory procedures

At baseline, clinical and demographic data were collected or abstracted from electronic medical records. These included age, sex, weight, height, smoking status, alcohol use, living conditions, diabetes status and other known chronic illness, history of TB infection and isoniazid use. HIV rapid testing was performed in programmatic settings for those of unknown status as part of HIV prevention. Where feasible, a CD4 lymphocyte count was performed. Viral load testing was performed under programmatic settings per the existing national guidelines. All participants on ART for at least 6 months had viral load testing done at the same time as the QFT-Plus test. All participants that had been on ART for less than 6 months at the time of the QFT-Plus test had viral load done after 6 months of ART use according to national guidelines. The ART regimen and initiation date were collected, and the date was used to determine the duration of ART use. Viral load level (≤ 40 versus > 40 copies/ml) and ART duration (≤3 versus >3 years) were used as a proxy for immune reconstitution and competency in the absence of CD4 lymphocyte count.

We used the QuantiFERON-TB Gold Plus (QFT-Plus) assay for TB infection diagnosis according to the manufacturer’s protocol. [29] Briefly, six ml of peripheral venous whole blood was collected into lithium heparin vacutainers and transported to the laboratory within two hours of collection. One ml was transferred into the four QFT-Plus blood collection tubes, the Nil tube, TB1 tube, TB2 tube, and the mitogen tube within 8 hours of collection and incubated at 37 °C for 16 hours. The tubes were centrifuged for 15 minutes at 2000 to 3000 x g RCF (g). Plasma was collected and assessed using the standard ELISA.

The results were interpreted according to the manufacturer’s instructions. [29] A QFT-Plus result was interpreted as positive when the IFN-γ response to one or both MTB-specific antigens was ≥0.35 IU/ml and ≥ 25% of Nil value, irrespective of the IFN-γ response to the mitogen control. A QFT-Plus result was interpreted as negative when the IFN-γ response to both MTB-specific proteins was <0.35 IU/ml, or ≥ 0.35 IU/ml and <25% of Nil value, with a response to the mitogen control ≥0.5 IU/ml. A QFT-Plus result was interpreted as indeterminate when the IFN-γ response to both MTB-specific proteins was <0.35 IU/ml, or ≥ 0.35 IU/ml and <25% of Nil value, with a response to the mitogen control <0.5 IU/ml. A QFT-Plus result was interpreted as indeterminate when the IFN-γ response was above the cut-off in the nil control, irrespective of the IFN-γ response to the MTB-specific antigens and the mitogen control.[29]

### Sample size

The sample size was determined based on an estimated TB infection prevalence of 35% in exposed [32] and 10% in unexposed, [10] at a 95% level of confidence and 80% power. Thus, a sample of 102 with continuity correction was sufficient, and all 121 eligible participants that were HIV-infected and that consented were selected for the study.

### Statistical analysis

After appropriate data cleaning, the analysis was performed using Stata Statistical Software version 17 (StataCorp LP, College Station, TX, USA). Descriptive statistics were used to summarise the sociodemographic and clinical characteristics of the participants. Pearson’s chi-square and Fisher’s exact tests were used to test the association between categorical variables and outcomes, and the odds ratios were reported. For continuous variables, the Student’s t-test was used. The multivariate logistic regression analyses included factors with a p-value < 0.10. Statistical significance was set at p-value <0.05. Missing data were addressed by removal to reduce the chance of biased estimates. Specifically, CD4 lymphocyte count data that were missing completely at random were excluded from the analysis and substituted with the duration of ART use. Complete case analysis was performed during multivariate regression (Figure 1).

### Patient and public involvement

It was not possible to involve patients or the public in the design, conduct, reporting, or dissemination plans of our research. Patients were involved to the extent to which they provided informed consent and allowed for sample collection for laboratory testing. The study results will be communicated to willing participants and community members.

### Ethical considerations

This study was approved by the Kenyatta National Hospital and the University of Nairobi Institutional Review Board.

## RESULTS

### Socio-demographic and clinical characteristics of participants

Among the 121 study participants living with HIV included in this analysis, females were the majority at 74.4% (90/121). The mean age of all participants was 38.4 years with a standard deviation (SD) of 10.8, and 51.2% (62/121) of the participants were ≥40 years. By BMI, 15.7% (19/121) of the participants were categorised as obese, 30.6% (37/121) as overweight, 49.6% (60/121) as normal, and 4.1% (5/121) as underweight. Among the social risk factors, 18.2% (22/121) were cigarette smokers, 20.7% (25/121) used alcohol, 27.3% (33/121) reported living in a crowded place, and 26.4% (32/121) had known contact with TB cases. Regarding clinical and laboratory characteristics, only 2.5% (3/121) had diabetes, and 38.1% (45 of 118 with complete ART duration data) had been on ART for >3 years. All participants were on the recommended first-line regimens with a backbone of tenofovir and lamivudine, combined with dolutegravir in 95% (115) and efavirenz in the remaining 5%. The participants with viral load <40 copies per ml were 85.7% (102 of 119 with complete viral load data). Table 1 shows the participants’ sociodemographic and clinical characteristics.

**Table 1.**
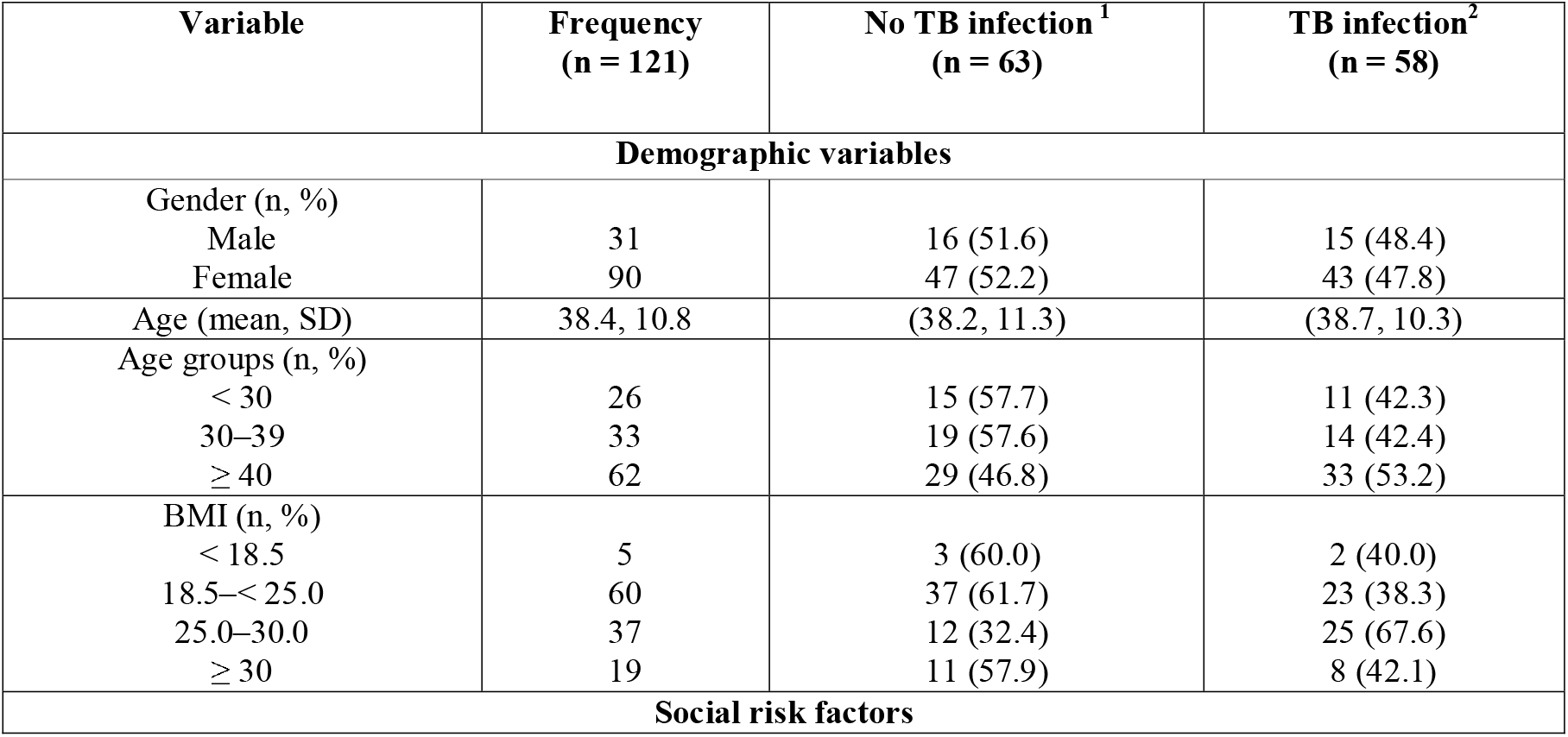
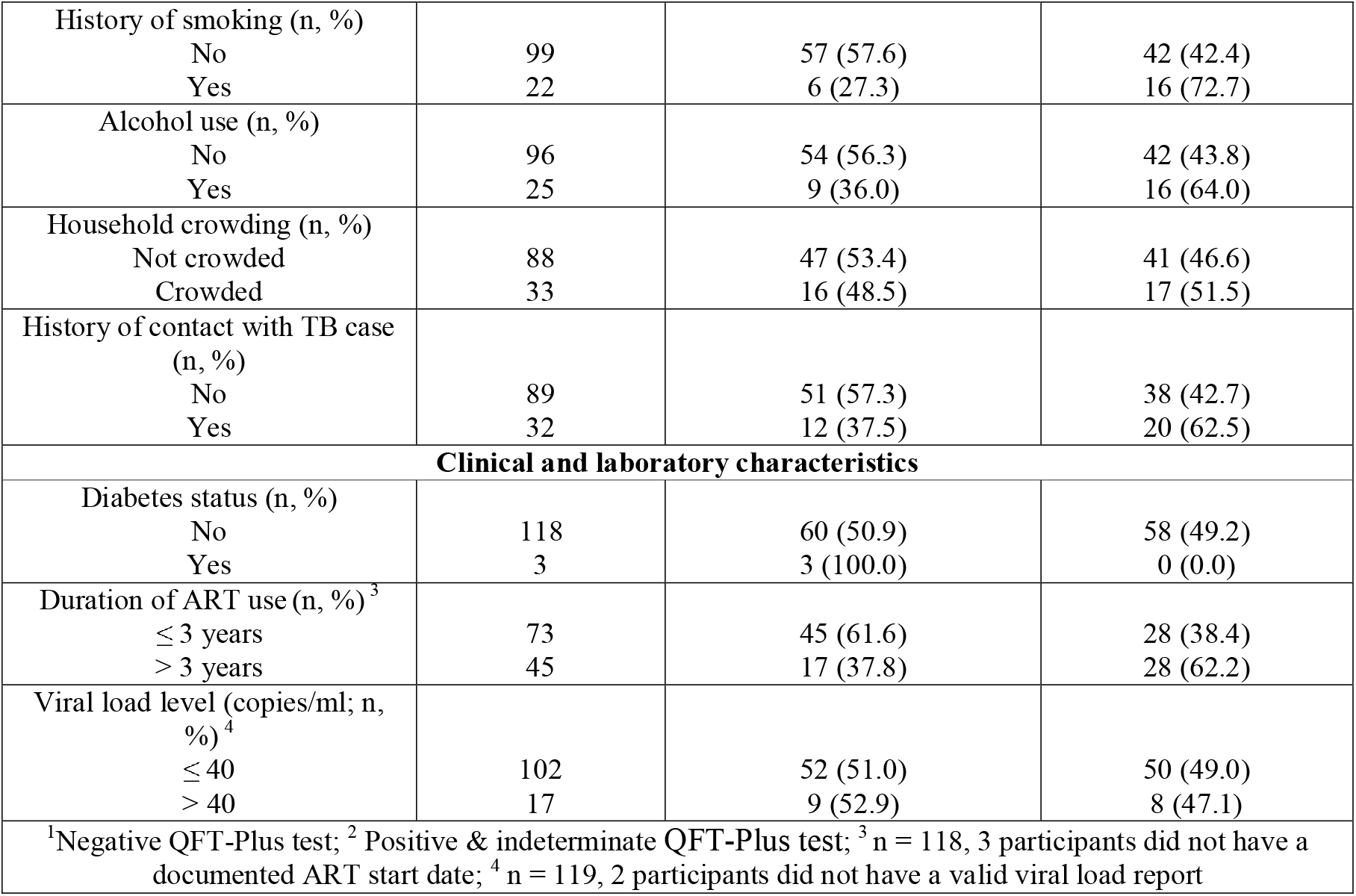
Socio-demographic and clinical characteristics of participants (n 121)

| Variable | Frequency<br>(n = 121) | No TB infection<sup>1</sup><br>(n = 63) | TB infection<sup>2</sup><br>(n = 58) |
| --- | --- | --- | --- |
| **Demographic variables** |  |  |  |
| Gender (n, %) |  |  |  |
| Male | 31 | 16 (51.6) | 15 (48.4) |
| Female | 90 | 47 (52.2) | 43 (47.8) |
| Age (mean, SD) | 38.4, 10.8 | (38.2, 11.3) | (38.7, 10.3) |
| Age groups (n, %) |  |  |  |
| $< 30$ | 26 | 15 (57.7) | 11 (42.3) |
| 30–39 | 33 | 19 (57.6) | 14 (42.4) |
| $\geq 40$ | 62 | 29 (46.8) | 33 (53.2) |
| BMI (n, %) |  |  |  |
| $< 18.5$ | 5 | 3 (60.0) | 2 (40.0) |
| 18.5–$< 25.0$ | 60 | 37 (61.7) | 23 (38.3) |
| 25.0–30.0 | 37 | 12 (32.4) | 25 (67.6) |
| $\geq 30$ | 19 | 11 (57.9) | 8 (42.1) |
| **Social risk factors** |  |  |  |
| History of smoking (n, %) |  |  |  |
| No | 99 | 57 (57.6) | 42 (42.4) |
| Yes | 22 | 6 (27.3) | 16 (72.7) |
| Alcohol use (n, %) |  |  |  |
| No | 96 | 54 (56.3) | 42 (43.8) |
| Yes | 25 | 9 (36.0) | 16 (64.0) |
| Household crowding (n, %) |  |  |  |
| Not crowded | 88 | 47 (53.4) | 41 (46.6) |
| Crowded | 33 | 16 (48.5) | 17 (51.5) |
| History of contact with TB case (n, %) |  |  |  |
| No | 89 | 51 (57.3) | 38 (42.7) |
| Yes | 32 | 12 (37.5) | 20 (62.5) |
| **Clinical and laboratory characteristics** |  |  |  |
| Diabetes status (n, %) |  |  |  |
| No | 118 | 60 (50.9) | 58 (49.2) |
| Yes | 3 | 3 (100.0) | 0 (0.0) |
| Duration of ART use (n, %) <sup>3</sup> |  |  |  |
| ≤ 3 years | 73 | 45 (61.6) | 28 (38.4) |
| > 3 years | 45 | 17 (37.8) | 28 (62.2) |
| Viral load level (copies/ml; n, %) <sup>4</sup> |  |  |  |
| ≤ 40 | 102 | 52 (51.0) | 50 (49.0) |
| > 40 | 17 | 9 (52.9) | 8 (47.1) |
| <sup>1</sup>Negative QFT-Plus test; <sup>2</sup> Positive & indeterminate QFT-Plus test; <sup>3</sup> n = 118, 3 participants did not have a documented ART start date; <sup>4</sup> n = 119, 2 participants did not have a valid viral load report |  |  |  |

### Prevalence of TB infection estimated by QFT-Plus test results

The prevalence of TB infection estimated by QFT-Plus test results in this population of PLHIV was 47.9% (58/121; Table 1). This comprised 39.7% (48/121) with a positive QFT-Plus test report and 8.3% (10/121) with an indeterminate QFT-Plus test report. Participants with an indeterminate QFT-Plus are described in Table 2. Among the participants with indeterminate results, 60% (6/10) were female, and 80% (8/10) were ≥30 years old. All the participants reported having HIV as the only known underlying chronic illness. Only 30% (3/10) reported cigarette use, and only 40% (4/10) reported alcohol use. Sixty per cent (6/10) of participants had been on ART for less than one year. In eight of the ten participants with indeterminate results, the interferon-gamma (IFN-γ) level in response to mitogen was below the cut-off of 0.5 IU/mL for Mitogen minus Nil (Table 2). Three of the ten participants had the IFN-γ response in the Nil tube above the cut-off of 8.0 IU/mL (Table 2), signifying an ongoing immune response.

**Table 2.**
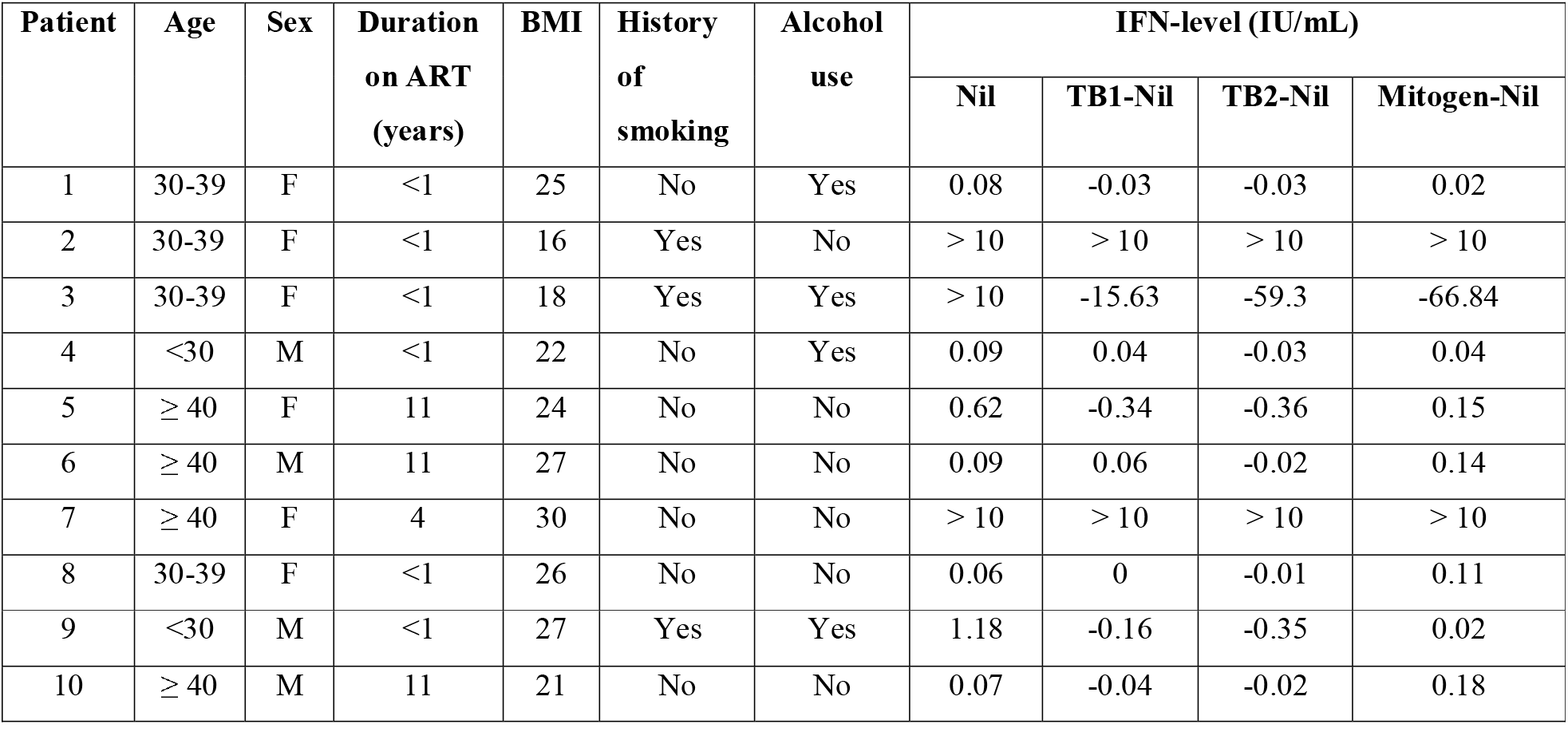
Characteristics of participants with indeterminate QFT-Plus test results (n=10)

### Determinants of TB infection estimated by QFT-Plus test results

The mean age of participants with TB infection was 38.7 years (standard deviation [SD] 10.3) compared to 38.2 years (SD 11.3) among those without TB infection (*p*=0.6022) (Table 3). On bivariate analysis, increased odds of having TB infection were observed among participants aged ≥40 years (*p*=0.353, odds ratio [OR] 1.55, 95% confidence interval [CI] 0.61–3.95), those with reported alcohol use (*p*=0.071, OR 2.29, 95% CI 0.90–5.78), those living in a crowded place (*p*=0.631, OR 1.22, 95% CI 0.54–2.72), and those with history of contact with a case of TB (*p*=0.056, OR 2.24, 95% CI 0.96–5.21). However, these findings were not statistically significant at *p* <0.05 (Table 3).

**Table 3.**
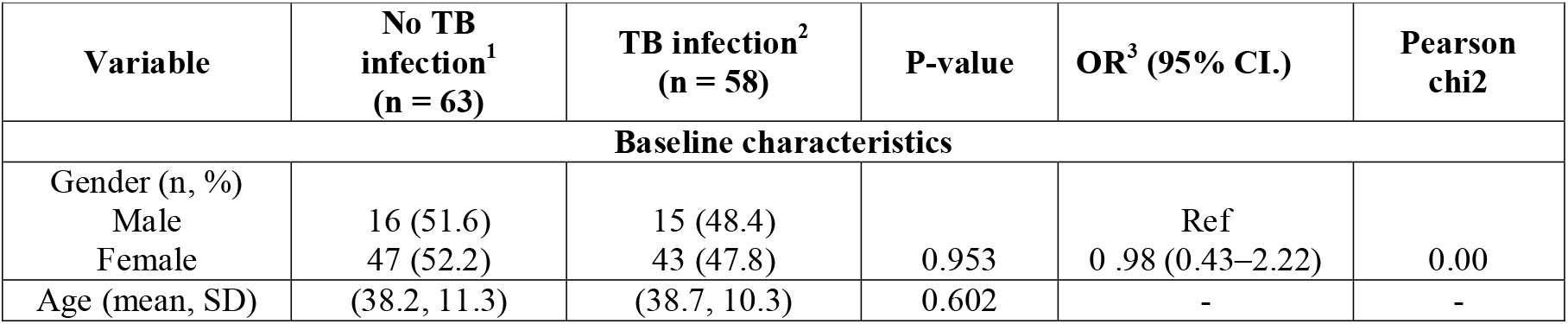
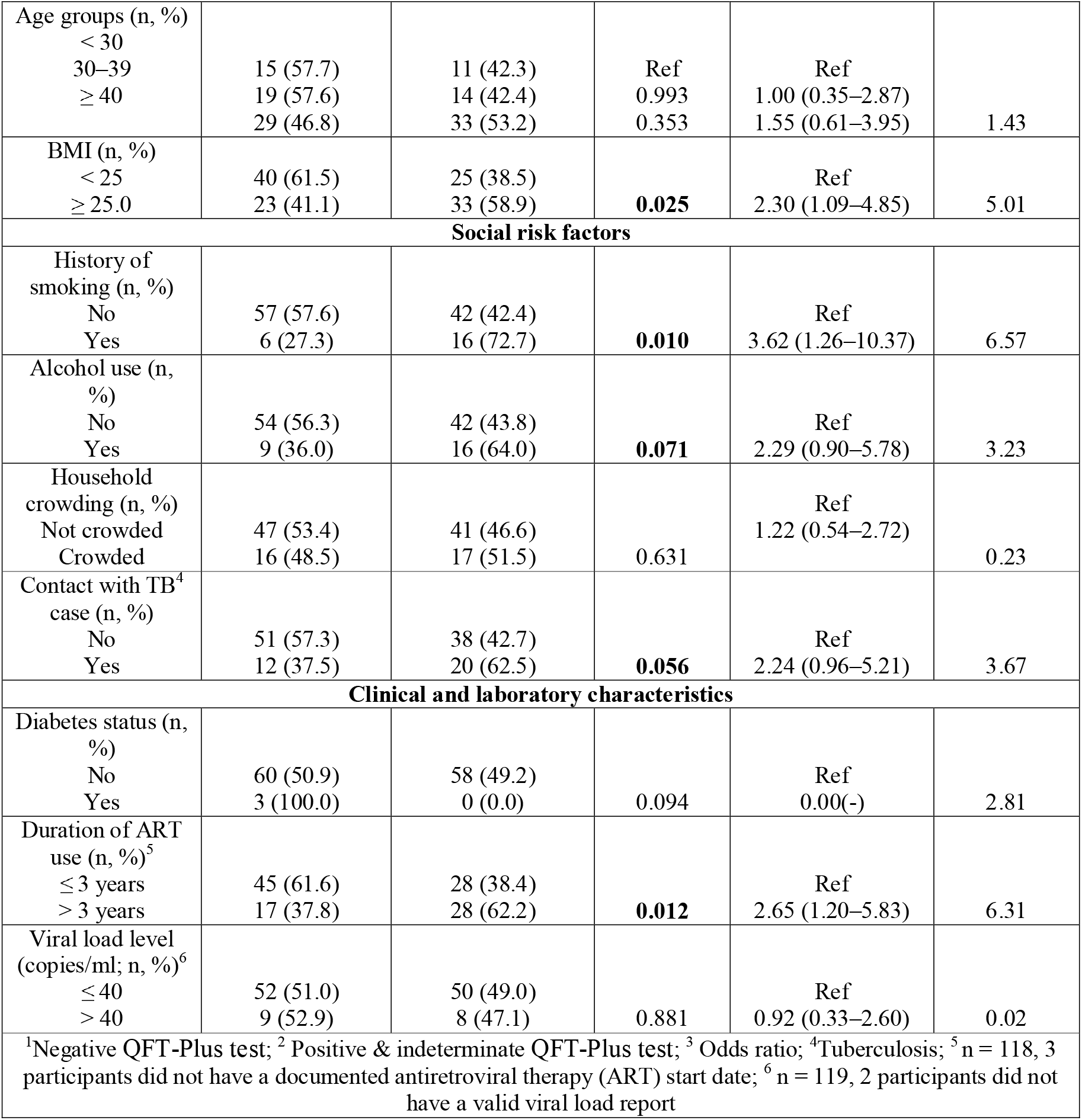
Determinants of TB infection among PLHIV – bivariate analysis (n 121)

| Variable | No TB infection<sup>1</sup> (n = 63) | TB infection<sup>2</sup> (n = 58) | P-value | OR<sup>3</sup> (95% CI.) | Pearson chi<sup>2</sup> |
| --- | --- | --- | --- | --- | --- |
| **Baseline characteristics** |  |  |  |  |  |
| Gender (n, %) |  |  |  |  |  |
| Male | 16 (51.6) | 15 (48.4) |  | Ref |  |
| Female | 47 (52.2) | 43 (47.8) | 0.953 | 0.98 (0.43–2.22) | 0.00 |
| Age (mean, SD) | (38.2, 11.3) | (38.7, 10.3) | 0.602 | - | - |
| Age groups (n, %) |  |  |  |  |  |
| < 30 |  |  |  |  |  |
| 30–39 | 15 (57.7) | 11 (42.3) | Ref | Ref |  |
| ≥ 40 | 19 (57.6) | 14 (42.4) | 0.993 | 1.00 (0.35–2.87) |  |
|  | 29 (46.8) | 33 (53.2) | 0.353 | 1.55 (0.61–3.95) | 1.43 |
| BMI (n, %) |  |  |  |  |  |
| < 25 | 40 (61.5) | 25 (38.5) |  | Ref |  |
| ≥ 25.0 | 23 (41.1) | 33 (58.9) | **0.025** | 2.30 (1.09–4.85) | 5.01 |
| **Social risk factors** |  |  |  |  |  |
| History of smoking (n, %) |  |  |  |  |  |
| No | 57 (57.6) | 42 (42.4) |  | Ref |  |
| Yes | 6 (27.3) | 16 (72.7) | **0.010** | 3.62 (1.26–10.37) | 6.57 |
| Alcohol use (n, %) |  |  |  |  |  |
| No | 54 (56.3) | 42 (43.8) |  | Ref |  |
| Yes | 9 (36.0) | 16 (64.0) | **0.071** | 2.29 (0.90–5.78) | 3.23 |
| Household crowding (n, %) |  |  |  |  |  |
| Not crowded | 47 (53.4) | 41 (46.6) |  | Ref |  |
| Crowded | 16 (48.5) | 17 (51.5) | 0.631 | 1.22 (0.54–2.72) | 0.23 |
| Contact with TB<sup>4</sup> case (n, %) |  |  |  |  |  |
| No | 51 (57.3) | 38 (42.7) |  | Ref |  |
| Yes | 12 (37.5) | 20 (62.5) | **0.056** | 2.24 (0.96–5.21) | 3.67 |
| **Clinical and laboratory characteristics** |  |  |  |  |  |
| Diabetes status (n, %) |  |  |  |  |  |
| No | 60 (50.9) | 58 (49.2) |  | Ref |  |
| Yes | 3 (100.0) | 0 (0.0) | 0.094 | 0.00(-) | 2.81 |
| Duration of ART use (n, %)<sup>5</sup> |  |  |  |  |  |
| ≤ 3 years | 45 (61.6) | 28 (38.4) |  | Ref |  |
| > 3 years | 17 (37.8) | 28 (62.2) | **0.012** | 2.65 (1.20–5.83) | 6.31 |
| Viral load level (copies/ml; n, %)<sup>6</sup> |  |  |  |  |  |
| ≤ 40 | 52 (51.0) | 50 (49.0) |  | Ref |  |
| > 40 | 9 (52.9) | 8 (47.1) | 0.881 | 0.92 (0.33–2.60) | 0.02 |
| <sup>1</sup>Negative QFT-Plus test; <sup>2</sup> Positive & indeterminate QFT-Plus test; <sup>3</sup> Odds ratio; <sup>4</sup>Tuberculosis; <sup>5</sup> n = 118, 3 participants did not have a documented antiretroviral therapy (ART) start date; <sup>6</sup> n = 119, 2 participants did not have a valid viral load report |  |  |  |  |  |

BMI (*p*=0.025, OR 2.30, 95% CI 1.09–4.85), a history of smoking cigarettes (*p*=0.010, OR 3.62, 95% CI 1.26–10.37), and duration of ART use >3 years (*p*=0.012, OR 2.65, 95% CI 1.20–5.83) were significantly associated with TB infection (Table 3). Being > 40 years old (*p*=0.005, OR 4.57, 95% CI 1.42-14.7) and having a positive QFT-Plus test (*p*=0.009, OR 2.89, 95% CI 1.25-6.65) were associated with being on ART > 3 years. Although increased odds of being on ART > 3 years were observed among participants with indeterminate results (*p*=0.420, OR 1.76, 95% CI 0.44-7.15), this finding was not statistically significant. Reduced odds of being on ART > 3 years were observed among participants having a viral load of >40 copies/ml (*p*=0.772, OR 0.85, 95% CI 0.29-2.51).

On multivariate analysis including factors with *p*<0.10 (Table 4), being obese/overweight (BMI ≥25; p=0.013, adjusted odds ratio (aOR) 2.90, 95% CI 1.25–6.74) and being on ART for >3 years (*p*=0.013, aOR 3.99, 95% CI 1.55–10.28) were independently associated with TB infection. There were increased odds of TB infection among those using alcohol (*p*=0.066, aOR 2.88, 95% CI 0.93– 8.91) (Table 4).

**Table 4.** Logistic regression analysis for factors associated with TB infection (n=118)

| Variable | No TB infection<sup>1</sup><br>(n = 62) | TB infection<sup>2</sup><br>(n = 56) | P-value | cOR<sup>3</sup> (95% CI.) | aOR<sup>4</sup> (95% CI.) |
| --- | --- | --- | --- | --- | --- |
| **Baseline characteristics** |  |  |  |  |  |
| BMI (n, %) |  |  |  |  |  |
| 18.5–< 25 | 39 (61.9) | 24 (38.1) | **0.013** | Ref | Ref |
| ≥ 25.0 | 23 (41.8) | 32 (58.2) |  | 2.26 (1.08–4.73) | 2.90 (1.25–6.74) |
| **Social risk factors** |  |  |  |  |  |
| History of smoking (n, %) |  |  |  |  |  |
| No | 56 (57.7) | 41 (42.3) | 0.253 | Ref | Ref |
| Yes | 6 (28.6) | 15 (71.4) |  | 3.41 (1.22–9.55) | 1.72 (0.44–6.67) |
| Alcohol use (n, %) |  |  |  |  |  |
| No | 53 (57.0) | 40 (43.0) | 0.066 | Ref | Ref |
| Yes | 9 (36.0) | 16 (64.0) |  | 2.36 (0.94–5.88) | 2.88 (0.93–8.91) |
| Contact with TB<sup>5</sup><br>case (n, %)<br>No<br>Yes | 51 (57.3)<br>11 (37.9) | 38 (42.7)<br>18 (62.1) | 0.367 | Ref<br>2.20 (0.93–5.19) | Ref<br>1.69 (0.54–5.33) |
| **Clinical and laboratory characteristics** |  |  |  |  |  |
| Duration of ART<br>use (n, %)<br>≤ 3 years<br>> 3 years | 45 (61.6)<br>17 (37.8) | 28 (38.4)<br>28 (62.2) | Ref<br>**0.013** | Ref<br>2.65 (1.23–5.69) | Ref<br>3.99 (1.55–10.28) |
| <sup>1</sup>Negative QFT-Plus test; <sup>2</sup>Positive & indeterminate QFT-Plus test; <sup>3</sup>crude odds ratio; <sup>4</sup>adjusted odds ratio; <sup>5</sup>Tuberculosis |  |  |  |  |  |

## DISCUSSION

We used the QFT-Plus test to estimate the prevalence and determinants of TB infection among PLHIV in the context of a high TB/HIV burden, near-universal ART access, and widespread TPT implementation. We found that the QFT-Plus test positivity in this population of PLHIV was 39.7%. This rate corresponds to estimates from the general population in an equivalent context. [33] This differs slightly from a similar study conducted among pregnant women in Kenya, where 35.8% of the pregnant women living with HIV had TB infection by QFT-Plus test positivity rate equivalent to those without HIV. [32] In Nigeria, in a similar high HIV/TB burden setup, Aladesanmi et al. found that 53% of PLHIV had TB infection. [34] Globally, TB infection is estimated at 25% of the world population. [1, 2] This variability, both geographically and by population, underscores the need for regular context-specific determination of the prevalence of TB infection for policy-making and service delivery cost allocation.

The most significant benefit following TPT is among PLHIV with confirmed TB infection. [17] Further, a recent review demonstrated higher proportions of PLHIV starting and completing TPT with testing for TB infection. [35] The current WHO and local guidelines indicate that confirming TB infection is not a prerequisite for TPT among PLHIV. [9, 18-20, 28] It is essential to consider the clinical and cost implications of treating individuals who may not benefit, either because they’re uninfected or because of non-adherence to TPT. [17, 35] From our findings, approximately 60% of PLHIV receiving TPT may not benefit, potentially exposing them to avoidable adverse drug events, drug-drug interactions, polypharmacy, and increased risk of non-adherence. Although a recent review concluded that providing TPT to PLHIV is cost-effective for preventing TB disease, [36] only two studies assessing the incremental cost of test-directed treatment from lower and middle-income countries were included. [37, 38] Furthermore, these were conducted among pregnant women, and the findings may not be generalisable. This, therefore, remains an area for further investigation.

We found that indeterminate results were reported in 8.3% of the cases, a rate similar to that of the general population but lower than those reported among PLHIV. [39-41] An altered immune response increases the likelihood of indeterminate results. [13, 39, 40, 42-44] In HIV, a low CD4 cell count is often implicated. [41, 45, 46] However, HIV can lead to indeterminate results even with normal CD4 cell counts due to altered T-cell function. [47-49] The rate of indeterminate result findings comparable to that of the average population in our study can be explained in two ways. First, although CD4 counts were unavailable, most participants-initiated ART early in the ‘Test and Treat’ era. We speculated that most had competent immunity. [50] Second, most participants had been on ART long enough (median duration on ART being 3.9 years), and ART duration influences immune recovery. [50-53]

A QFT-Plus test results is interpreted as indeterminate when there is a decrease in IFN-γ production in the mitogen tube (positive control) or/and an increase in the Nil tube (negative control). [29] In our study, we found decreased production of IFN-γ in the mitogen tube in 8 out of 10 cases. An altered immune response will lead to an insufficient reaction in the mitogen tube, corresponding to the incapacity of lymphocytes to secrete IFN-γ. [46, 54-58] Possibly, the T lymphocytes among the participants with indeterminate results were compromised in quality and were thus unable to produce sufficient levels of IFN-γ. [46, 57] In support, most of the participants with indeterminate results had been on ART for less than a year, all were ambulatory, and none had a known illness other than HIV. In cases of immune suppression, repeat testing is advised after recovery. [54, 55] Further, with indeterminate results, TB infection cannot be ruled out with certainty, and the prognosis may be poorer. [54, 55] The increased response in the Nil tube found in three participants may signify residual IFN-γ due to ongoing infection, [58] and TB infection could not be ruled out.

Interpreting indeterminate results is a clinical dilemma. The WHO and local Kenyan guidelines do not require testing for TB infection before initiating TPT, and there’s no guidance for handling indeterminate results where testing is done. [9, 18, 20, 28] Given the clinical implication and potential benefit of TPT in PLHIV [30, 31], the possibility of immune suppression among participants with indeterminate results, the uncertainty around prognostic implications of indeterminate results [54, 55] and the resource limitation that precluded repeat testing during follow-up, the operational definition of TB infection used in the present study was justified. Indeterminate results were included in the definition of TB infection, bringing the proportion of PLHIV classified as having TB infection to 47.9%.

TB infection was significantly higher among participants who were obese/overweight (BMI≥25) and who had been on ART for >3 years. The association with obesity differed from previous findings, although PLHIV were not explicitly considered in these studies. [59-61] Obesity is associated with an increased risk of diabetes, [61] which increases the risk of TB infection and progression to active TB. [62, 63] Additionally, ART is implicated in various components of metabolic syndrome, including an increased risk of hyperglycaemia and diabetes.[41] However, our study only assessed self-reported diabetes and did not measure the haemoglobin A1c (HbA1c) level to determine its association with TB infection. We hypothesised that being on ART for >3 years resulted in CD4 recovery to at least the lower end of the normal range with increased yield from QFT-Plus test testing. [46] There was a non-significant association of viral load with ART duration. Since the viral load was not done at the point of QFT-Plus testing, it was not suitable to assess immune reconstitution, explaining our findings. Although the age category was not associated with TB infection, [14, 64, 65] those on ART longer were also older. Longer cumulative exposure time that is expected with increasing age could be a source of residual confounding. Compared to those without TB infection, those on ART longer had higher odds of being indeterminate. This is contrary to what we would expect; however, the study was not sufficiently powered to assess this association, given that the numbers with indeterminate results were small.

Alcohol use increases the likelihood of TB infection and disease. [66, 67] Although not significantly associated with TB infection on multivariable analysis, there were increased odds of TB infection with a trend towards significance. Structurally, people who consume alcohol are more likely to congregate in crowded places with high TB transmission rates. Alcohol is also known to cause immune suppression, which increases the risk of infection. [67] There was no association between TB infection and living in a crowded place or contact with a person with TB compared to findings from other studies. [33] This could be due to the small sample size and the reduced power required to detect an association. Additionally, household crowding and the history of contact with a TB case were measured by self-report. This is subjective and disposed to social desirability and recall bias.

## STUDY STRENGTHS AND LIMITATIONS

Our study had several strengths. The involvement of participants visiting the national referral hospital, who were a cosmopolitan group, increased the ability to generalise our findings. We excluded participants with prior exposure to TPT, thus reducing the chance of outcome misclassification. We considered the clinical implications of an indeterminate QFT-Plus results for a resource-limited setup. A limitation of our study is that we did not collect data on CD4 counts and could not account for the association of immune status with TB infection. This was due to changes in the guidelines, precluding the use of CD4 for HIV monitoring. Since the viral load was not done at the point of QFT-Plus testing, there was a misclassification bias. We used years of ART as a proxy for immune recovery and competency to address this limitation.

Second, alcohol use, cigarette use, household crowding, history of contact with a TB case and presence of other chronic conditions, including autoimmune diseases, were based on self-reporting. This is subject to error and social desirability, and recall bias. Third, obesity was not correlated with other metabolic dysfunctions, and diabetes status was not objectively ascertained. Finally, this was a cross-sectional study, and no long-term follow-up was conducted to see those who developed TB disease. A larger sample size would have had more power to detect associations. Therefore, studies with larger sample sizes are necessary to elucidate better the factors affecting indeterminate QFT-plus tests.

## Data Availability

All data produced in the present study are available upon reasonable request to the authors

## CONCLUSION AND RECOMMENDATION

The prevalence of TB infection in this population of PLHIV who had not received TPT is higher than the global estimate, depicting high levels of exposure and better performance of the QFT-Plus test with early and longer ART use. The rates of indeterminate results were the same as those reported for the general population, further supporting the effect of ART on immunity and QFT-Plus test sensitivity. Given the existing evidence, it is important to consider the clinical and cost implications of test-directed TPT for PLHIV in low and middle-income countries. This will better inform policies towards recommending test-directed TPT in PLHIV never exposed to TPT. Clear directives on handling indeterminate QFT-Plus test results in PLHIV are needed. People with vulnerabilities, such as obesity and alcoholism, need to be targeted for TPT. A more in-depth analysis of the determinants of TB infection using a larger sample size is recommended.

### ABBREVIATIONS

ART: antiretroviral therapy
CD4: cluster of differentiation 4
CI: confidence interval
HIV: human immunodeficiency virus
HBC: high burden country
IGRA: interferon-gamma release assay
LTBI: latent tuberculosis infection
PLHIV: people living with human immunodeficiency virus
SD: standard deviation
TB: tuberculosis
TPT: tuberculosis preventive therapy
WHO: World Health Organization

## DECLARATIONS

### Ethics approval and consent to participate

This study was approved by the Kenyatta National Hospital/University of Nairobi Institutional Review Board (KNH/UON ERB) under Ref. KNH-ERC/A/375.

### Consent for publication

No individual personal data are included in this manuscript. All data were anonymised and extracted after obtaining informed consent and ethical approval to conduct the study.

### Availability of data and materials

All data generated or analysed during this study are not publicly available due to privacy policy regulations but are available from the corresponding author upon reasonable request.

### Author contributions

Concept development and study design: LNN, JOM, VN. Supervision of the Study: LNN, JOM, VN, and MM. Data analysis: LNN and VN. Critically revised manuscript: LNN, JOM, VN, and MM. All authors have read and approved the final draft for publication.

## Acknowledgements

LNN was supported by CARTA. CARTA is jointly led by the African Population and Health Research Centre and the University of the Witwatersrand and funded by the Carnegie Corporation of New York (Grant No. G-19–57145), Sida (Grant No:54100113), Uppsala Monitoring Centre, Norwegian Agency for Development Cooperation (Norad), the Wellcome Trust (reference no. 107768/Z/15/Z), and the UK Foreign, Commonwealth & Development Office, with support from the Developing Excellence in Leadership, Training and Science in Africa (DELTAS Africa) programme. The statements made and views expressed are solely the responsibility of the Fellow. For open access, the authors have applied a CC BY public copyright licence to any Author Accepted Manuscript version arising from this submission.

LNN also received training from the NIH/Fogarty HIV Research Training Program (D43 TW011817 Tuberculosis & HIV Co-Infection Training Program in Kenya) during manuscript development.

We thank the patients who volunteered for this study and the research team involved in data collection. We also thank the staff and administration of the Kenyatta National Hospital’s comprehensive care clinic and Pumwani Maternity Hospital for supporting this work. We would also like to acknowledge the University of Nairobi, Faculty of Health Sciences, for doctoral training support.

We thank Editage (www.editage.com) for English language editing.

## Authors’ information

### Exclusive Licence

I, the Submitting Author has the right to grant and does grant on behalf of all authors of the Work (as defined in the below author licence), an exclusive licence and/or a non-exclusive licence for contributions from authors where BMJ has agreed a CC-BY licence shall apply; on a worldwide, perpetual, irrevocable, royalty-free basis to BMJ Publishing Group Ltd (“BMJ”) its licensees and where the relevant Journal is co-owned by BMJ to the co-owners of the Journal, to publish the Work in BMJ Open Respiratory Research and any other BMJ products and to exploit all rights, as set out in our <u>licence</u>.

## References

1. Dye C, Scheele S, Dolin P, et al. Global burden of tuberculosis. JAMA: the journal of the American Medical Association. 1999;282:677–86.

2. Dye C, Scheele S, Dolin P, et al. Consensus statement. Global burden of tuberculosis: estimated incidence, prevalence, and mortality by country. WHO Global Surveillance and Monitoring Project. Jama. 1999;282(7):677–86.

3. World Health Organization. Global tuberculosis report. Geneva; 2022. https://www.who.int/teams/global-tuberculosis-programme/tb-reports/global-tuberculosis-report-2022 (accessed 18 Jan 2023).

4. Ford N MA, Shubber Z, et al. TB as a cause of hospitalization and in-hospital mortality among people living with HIV worldwide: a systematic review and meta-analysis. Journal of the International AIDS Society. 2016;19(1):20714.

5. AIDS info (website). Geneva: UNAIDS; 2022 (https://aidsinfo.unaids.org/). (accessed 18 Jan 2023).

6. World Health Organisation. Tuberculosis profile: Global (website). 2021 https://worldhealthorg.shinyapps.io/tb_profiles/?_inputs_&lan=%22EN%22&entity_type=%22group%22&group_code=%22global%22 (accessed 24 Jan 2023) [

7. Ai JW, Ruan QL, Liu QH, et al. Updates on the risk factors for latent tuberculosis reactivation and their managements. Emerg Microbes Infect. 2016;5(2):e10.

8. Vynnycky E, Fine PE. The natural history of tuberculosis: the implications of age-dependent risks of disease and the role of reinfection. Epidemiology and infection. 1997;119(2):183–201.

9. World Health Organisation. Consolidated guidelines on tuberculosis. Module 1: prevention – tuberculosis preventive treatment. Geneva: World Health Organization; 2020. https://www.who.int/publications/i/item/9789240001503 (accessed 13 July 2022).

10. Houben RMGJ, Dodd PJ. The Global Burden of Latent Tuberculosis Infection: A Re-estimation Using Mathematical Modelling. PLOS Medicine. 2016;13(10):e1002152.

11. Cohen A, Mathiasen VD, Schön T, et al. The global prevalence of latent tuberculosis: a systematic review and meta-analysis. Eur Respir J. 2019;54(3).

12. Basera TJ, Ncayiyana J, Engel ME. Prevalence and risk factors of latent tuberculosis infection in Africa: a systematic review and meta-analysis protocol. BMJ Open. 2017;7(7):e012636.

13. González-Moreno J, García-Gasalla M, Losada-López I, et al. IGRA testing in patients with immune-mediated inflammatory diseases: which factors influence the results? Rheumatol Int. 2018;38(2):267–73.

14. Ncayiyana JR, Bassett J, West N, et al. Prevalence of latent tuberculosis infection and predictive factors in an urban informal settlement in Johannesburg, South Africa: a cross-sectional study. BMC Infectious Diseases. 2016;16(1):661.

15. Santos JA, Duarte R, Nunes C. Host factors associated to false negative and indeterminate results in an interferon-γ release assay in patients with active tuberculosis. Pulmonology. 2020;26(6):353–62.

16. Antonucci G, Girardi E, Raviglione MC, et al. Risk Factors for Tuberculosis in HIV-lnfected Persons: A Prospective Cohort Study. Jama. 1995;274(2):143–8.

17. Akolo C, Adetifa I, Shepperd S, Volmink J. Treatment of latent tuberculosis infection in HIV infected persons. Cochrane Database of Systematic Reviews. 2010(1).

18. Ministry of Health, National AIDS & STI Control Program. Kenya HIV Prevention and Treatment Guidelines, 2022 Edition. Nairobi, Kenya: NASCOP, Aug 2022. Print.

19. World Health Organisation. Latent tuberculosis infection: updated and consolidated guidelines for programmatic management Geneva: World Health Organization; 2018 [updated 2018. https://apps.who.int/iris/handle/10665/260233 (accessed 13 July 2022).

20. Kenya Ministry of Health. Integrated guideline for tuberculosis, leprosy and lung disease. 2021. https://chskenya.org/wp-content/uploads/2022/04/integrated-guideline-for-tuberculosis-leprosy-and-lung-disease-2021.pdf (accessed 24 Jan 2023).

21. Joseph L, Gyorkos TW, Coupal L. Bayesian estimation of disease prevalence and the parameters of diagnostic tests in the absence of a gold standard. Am J Epidemiol. 1995;141(3):263–72.

22. Rogan WJ, Gladen B. Estimating prevalence from the results of a screening test. Am J Epidemiol. 1978;107(1):71–6.

23. Wolf T, Goetsch U, Oremek G, et al. Tuberculosis skin test, but not interferon-γ-releasing assays is affected by BCG vaccination in HIV patients. The Journal of infection. 2013;66(4):376–80.

24. Sultan B, Benn P, Mahungu T, et al. Comparison of two interferon-gamma release assays (QuantiFERON-TB Gold In-Tube and T-SPOT.TB) in testing for latent tuberculosis infection among HIV-infected adults. Int J STD AIDS. 2013;24(10):775–9.

25. Klautau GB, da Mota NVF, Salles MJC, et al. Interferon-γ release assay as a sensitive diagnostic tool of latent tuberculosis infection in patients with HIV: a cross-sectional study. BMC Infect Dis. 2018;18(1):585.

26. Shafeque A, Bigio J, Hogan CA, et al. Fourth-Generation QuantiFERON-TB Gold Plus: What Is the Evidence? Journal of clinical microbiology. 2020;58(9).

27. Xu Y, Yang Q, Zhou J, et al. Comparison of QuantiFERON-TB Gold In-Tube and QuantiFERON-TB Gold-Plus in the Diagnosis of Mycobacterium tuberculosis Infections in Immunocompromised Patients: a Real-World Study. Microbiol Spectr. 2022;10(2):e0187021.

28. Ministry of Health, National AIDS & STI Control Program. Guidelines on Use of Antiretroviral Drugs for Treating and Preventing HIV Infection in Kenya 2018 Edition. Nairobi, Kenya: NASCOP, August 2018. Print.

29. QuantiFERON-TB Gold Plus (QFT-Plus) ELISA Package Insert 02/2016. http://www.quantiferon.com/wp-content/uploads/2017/04/English_QFTPlus_ELISA_R04_022016.pdf (accessed Oct 2019).

30. Badje A, Moh R, Gabillard D et al. Effect of isoniazid preventive therapy on risk of death in west African, HIV-infected adults with high CD4 cell counts: long-term follow-up of the Temprano ANRS 12136 trial. Lancet Glob Health. 2017;5(11):e1080–e9.

31. The TEMPRANO ANRS 12136 Study Group. A Trial of Early Antiretrovirals and Isoniazid Preventive Therapy in Africa. N Engl J Med. 2015;373(9):808–22.

32. Kaplan SR, Escudero JN, Mecha J, et al. Interferon Gamma Release Assay and Tuberculin Skin Test Performance in Pregnant Women Living With and Without HIV. Journal of acquired immune deficiency syndromes (1999). 2022;89(1):98–107.

33. Jensen AV, Jensen L, Faurholt-Jepsen D, et al. The prevalence of latent Mycobacterium tuberculosis infection based on an interferon-γ release assay: a cross-sectional survey among urban adults in Mwanza, Tanzania. PloS one. 2013;8(5):e64008.

34. Aladesanmi AO, Ojuawo OB, Aladesanmi OO, et al. Diagnosis of latent tuberculosis among HIV infected patients in Ilorin, Nigeria using tuberculin skin test and interferon gamma release assay. Pan Afr Med J. 2021;38:24.

35. Bastos ML, Melnychuk L, Campbell JR, et al. The latent tuberculosis cascade-of-care among people living with HIV: A systematic review and meta-analysis. PLoS Med. 2021;18(9):e1003703.

36. Uppal A, Rahman S, Campbell JR, et al. Economic and modeling evidence for tuberculosis preventive therapy among people living with HIV: A systematic review and meta-analysis. PLOS Medicine. 2021;18(9):e1003712.

37. Kapoor S, Gupta A, Shah M. Cost-effectiveness of isoniazid preventive therapy for HIV-infected pregnant women in India. The international journal of tuberculosis and lung disease : the official journal of the International Union against Tuberculosis and Lung Disease. 2016;20(1):85–92.

38. Kim HY, Hanrahan CF, Martinson N, et al. Cost-effectiveness of universal isoniazid preventive therapy among HIV-infected pregnant women in South Africa. The international journal of tuberculosis and lung disease : the official journal of the International Union against Tuberculosis and Lung Disease. 2018;22(12):1435–42.

39. Oliveira S, Trajman A, Paniago AMM, et al. Frequency of indeterminate results from an interferon-gamma release assay among HIV-infected individuals. J Bras Pneumol. 2017;43(3):215–8.

40. Oni T, Gideon HP, Bangani N, et al. Risk factors associated with indeterminate gamma interferon responses in the assessment of latent tuberculosis infection in a high-incidence environment. Clinical and vaccine immunology : CVI. 2012;19(8):1243–7.

41. Luetkemeyer AF, Charlebois ED, Flores LL, et al. Comparison of an interferon-gamma release assay with tuberculin skin testing in HIV-infected individuals. Am J Respir Crit Care Med. 2007;175(7):737–42.

42. Jeong SJ, Han SH, Kim CO, et al. Predictive factors for indeterminate result on the QuantiFERON test in an intermediate tuberculosis-burden country. Journal of Infection. 2011;62(5):347–54.

43. Darby J, Black J, Buising K. Interferon-gamma release assays and the diagnosis of tuberculosis: have they found their place? Internal Medicine Journal. 2014;44(7):624–32.

44. Papay P, Eser A, Winkler S, et al. Predictors of indeterminate IFN-γ release assay in screening for latent TB in inflammatory bowel diseases. Eur J Clin Invest. 2011;41(10):1071–6.

45. Brock I, Ruhwald M, Lundgren B, et al. Latent tuberculosis in HIV positive, diagnosed by the M. tuberculosis specific interferon-gamma test. Respir Res. 2006;7(1):56.

46. Raby E, Moyo M, Devendra A, et al. The effects of HIV on the sensitivity of a whole blood IFN-gamma release assay in Zambian adults with active tuberculosis. PloS one. 2008;3(6):e2489.

47. Bhosale R, Alexander M, Deshpande P, et al. Stages of pregnancy and HIV affect diagnosis of tuberculosis infection and Mycobacterium tuberculosis (MTB)-induced immune response: Findings from PRACHITi, a cohort study in Pune, India. International journal of infectious diseases : IJID : official publication of the International Society for Infectious Diseases. 2021;112:205–11.

48. Kroon EE, Kinnear CJ, Orlova M F et al. An observational study identifying highly tuberculosis-exposed, HIV-1-positive but persistently TB, tuberculin and IGRA negative persons with M. tuberculosis specific antibodies in Cape Town, South Africa. EBioMedicine. 2020;61:103053.

49. Lawn SD, Bekker LG, Wood R. How effectively does HAART restore immune responses to Mycobacterium tuberculosis? Implications for tuberculosis control. AIDS (London, England). 2005;19(11):1113–24.

50. Le T, Wright EJ, Smith DM, et al. Enhanced CD4+ T-cell recovery with earlier HIV-1 antiretroviral therapy. N Engl J Med. 2013;368(3):218–30.

51. Kufa T, Shubber Z, MacLeod W, et al. CD4 count recovery and associated factors among individuals enrolled in the South African antiretroviral therapy programme: An analysis of national laboratory based data. PloS one. 2019;14(5):e0217742.

52. Fisk TL, Hon HM, Lennox JL, et al. Detection of latent tuberculosis among HIV-infected patients after initiation of highly active antiretroviral therapy. AIDS (London, England). 2003;17(7):1102–4.

53. Bishop J, DeShields S, Cunningham T, et al. CD4 Count Recovery After Antiretroviral Therapy Initiation in Patients Infected with the Human Immunodeficiency Virus. Am J of the Med Sci. 2016;352.

54. Sester M, van Leth F, Bruchfeld J, et al. Risk assessment of tuberculosis in immunocompromised patients. A TBNET study. Am J Respir Crit Care Med. 2014;190(10):1168–76.

55. Jacquier M, Binquet C, Manoha C, et al. Beyond QuantiFERON-TB Results, the Added Value of a Weak Mitogen Response. Front Med (Lausanne). 2022;9:876864.

56. Belliere J, Blancher A. QuantiFERON test interpretation in patients receiving immunosuppressive agents: an alert. Eur Respir J. 2017;49(4):1602102.

57. Aabye MG, Ravn P, PrayGod G J et al. The impact of HIV infection and CD4 cell count on the performance of an interferon gamma release assay in patients with pulmonary tuberculosis. PloS one. 2009;4(1):e4220.

58. Ariga H, Nagai H, Kurashima A, Hoshino Y, et al. Stratified Threshold Values of QuantiFERON Assay for Diagnosing Tuberculosis Infection in Immunocompromised Populations. Tuberc Res Treat. 2011;2011:940642.

59. Badawi A, Liu CJ. Obesity and Prevalence of Latent Tuberculosis: A Population-Based Survey. Infect Dis (Auckl). 2021;14:1178633721994607.

60. Critchley JA, Restrepo BI, Ronacher K, et al. Defining a Research Agenda to Address the Converging Epidemics of Tuberculosis and Diabetes: Part 1: Epidemiology and Clinical Management. Chest. 2017;152(1):165–73.

61. Roth J, Sahota N, Patel P, et al. Obesity paradox, obesity orthodox, and the metabolic syndrome: An approach to unity. Mol Med. 2017;22:873–85.

62. Barron MM, Shaw KM, Bullard KM, et al. Diabetes is associated with increased prevalence of latent tuberculosis infection: Findings from the National Health and Nutrition Examination Survey, 2011-2012. Diabetes Res Clin Pract. 2018;139:366–79.

63. Badawi A, Sayegh S, Sallam M, et al. The global relationship between the prevalence of diabetes mellitus and incidence of tuberculosis: 2000-2012. Glob J Health Sci. 2014;7(2):183–91.

64. Lin W-C, Lin H-H, Lee SS-J, et al. Prevalence of latent tuberculosis infection in persons with and without human immunodeficiency virus infection using two interferon-gamma release assays and tuberculin skin test in a low human immunodeficiency virus prevalence, intermediate tuberculosis-burden country. J Microbiol, Immunol Infect. 2016;49(5):729–36.

65. Kizza FN, List J, Nkwata AK, et al. Prevalence of latent tuberculosis infection and associated risk factors in an urban African setting. BMC Infect Dis. 2015;15:165.

66. Imtiaz S, Shield KD, Roerecke M, et al. Alcohol consumption as a risk factor for tuberculosis: meta-analyses and burden of disease. Eur Respir J. 2017;50(1).

67. Rehm J, Samokhvalov AV, Neuman MG, et al. The association between alcohol use, alcohol use disorders and tuberculosis (TB). A systematic review. BMC Public Health. 2009;9:450.

